# Large-scale reorganization of cortical hierarchy, dynamics, and information processing in subjective cognitive decline

**DOI:** 10.64898/2026.01.26.26344800

**Authors:** Xiaobo Liu, Xingyan Le, Zhengyuan Huang, Yuyin Wang, Qingbiao Zhang, Junbang Feng, Chuanming Li

**Affiliations:** Medical Imaging Department, Chongqing Emergency Medical Center, Chongqing University Central Hospital, School of Medicine, Chongqing University, Chongqing, China; McConnell Brain Imaging Centre, Montreal Neurological Institute, McGill University, Montreal, Canada; Department of Psychiatry, Renmin Hospital of Wuhan University, Wuhan, Hubei Province, China

**Keywords:** Subjective cognitive decline, preclinical Alzheimer’s disease, functional brain reorganization, hierarchical processing, neuroimaging biomarkers

## Abstract

Subjective cognitive decline (SCD) represents a clinically important but mechanistically elusive stage along the Alzheimer’s disease (AD) continuum, characterized by persistent cognitive complaints despite preserved performance on standard neuropsychological tests. Individuals with SCD show an increased risk of progression to mild cognitive impairment and dementia; however, the large-scale functional architecture underlying subjective cognitive symptoms remains poorly defined.

In this study, we investigated cortical functional hierarchy, hierarchical brain dynamics, and information-processing architecture in individuals with SCD using resting-state functional magnetic resonance imaging. We first characterized the principal functional gradient spanning sensory to transmodal association cortices and quantified its global dispersion. We then examined the directionality of hierarchical brain dynamics to assess alterations in bottom-up and top-down information flow. Finally, we applied an integrated information decomposition framework to quantify synergistic and redundant interactions across large-scale brain networks, and related these measures to clinical characteristics and molecular–cellular substrates.

Compared with healthy controls, individuals with SCD exhibited significant regional alterations in functional gradient organization, predominantly affecting association cortices, alongside increased gradient dispersion, indicating a less compact and more heterogeneous cortical hierarchy. Hierarchical dynamics were shifted toward enhanced bottom-up information flow, suggesting increased reliance on sensory-driven processing. Integrated information decomposition revealed widespread reductions in both synergistic and redundant interactions, reflecting impaired integrative capacity and reduced informational robustness of distributed brain systems. These functional alterations were associated with symptom duration and depressive burden, and showed spatial correspondence with neurotransmitter receptor distributions and cell-type–specific gene expression profiles.

Together, these findings demonstrate that SCD is characterized by coordinated disruptions in cortical hierarchy, hierarchical dynamics, and information-processing organization. Our results provide a systems-level account of early functional brain reorganization in SCD and support its conceptualization as an early functional manifestation of AD-related vulnerability, prior to objective cognitive impairment.

## Introduction

Subjective cognitive decline (SCD) is increasingly regarded as one of the earliest and most covert stages along the Alzheimer’s disease (AD) continuum ^1^. Individuals with SCD experience persistent cognitive complaints despite normal performance on standardized neuropsychological assessments, and show an elevated risk of progression to mild cognitive impairment and dementia compared with age-matched peers ^2^. The absence of reliable and reproducible objective biomarkers has limited early risk stratification and intervention, leaving the neural basis of subjective cognitive symptoms largely unresolved ^3^.

Accumulating evidence suggests that SCD does not merely reflect affective or psychological factors, but may index early reorganization of large-scale brain function that precedes overt cognitive impairment ^4^. In the healthy human brain, information processing is organized along a cortical hierarchy spanning from lower-order sensory and unimodal regions to higher-order transmodal association cortices ^5,6^. This hierarchical architecture supports the integration of sensory input with memory, internal representations, and abstract cognition ^7,8^. Association cortices are particularly vulnerable to aging and neurodegenerative processes due to their prolonged maturation, high metabolic demands, and dense long-range connectivity ^9,10^. Recent pioneering research demonstrates that atrophy in neurodegenerative diseases, including AD, systematically disrupts the functional hierarchy extending from primary sensory networks to high-level association areas ^11^. However, whether SCD is associated with systematic alterations in cortical hierarchical organization—and whether such alterations relate to symptom persistence and disease risk—remains unclear.

Recent advances in resting-state functional magnetic resonance imaging (rs-fMRI) have enabled the characterization of cortical organization in terms of continuous functional gradients^12,13^. The principal sensory-to-association gradient captures a fundamental axis of intrinsic brain organization, reflecting transitions from perceptual processing to higher-order cognitive integration ^14^. Functional gradients have proven sensitive to neurodevelopmental, psychiatric, and neurodegenerative disorders, including AD^15–17^. Yet it remains unknown whether disruptions of this hierarchical organization are already detectable at the SCD stage, or whether such changes manifest as increased dispersion of the cortical functional hierarchy.

More fundamentally, characterizing early functional reorganization requires moving beyond measures of connectivity strength to interrogate how information is processed and distributed across neural systems ^22^. Integrated information decomposition frameworks partition neural interactions into synergistic and redundant components, providing a principled account of distributed information processing ^23^. Synergistic interactions reflect integrative computations that emerge from the joint activity of multiple regions, whereas redundant interactions support robustness through shared information ^24,25^. Previous studies have shown that connectome neurodegeneration drives age-related changes in redundancy and synergy ^26^. Disruptions to these information-processing modes have been reported in neurological and psychiatric conditions, yet how synergy and redundancy are organized along the cortical hierarchy in SCD remains largely unexplored ^27^.

Here, we conceptualize SCD as a functional reorganization stage within the AD risk continuum and investigate large-scale cortical hierarchy, hierarchical dynamics, and information processing in individuals with SCD using resting-state fMRI. We first characterize alterations in the principal functional gradient and its global dispersion. We then examine hierarchical directionality of brain dynamics, assessing imbalances between bottom-up and top-down information flow. Finally, we apply integrated information decomposition to quantify changes in synergistic and redundant interactions across the cortex, and relate these functional features to clinical measures and underlying molecular and cellular substrates. By integrating spatial hierarchy, temporal dynamics, and mation-processing modes within a unified framework, this study aims to delineate the systems-level neural signature of SCD and to advance objective characterization of early AD risk.

## Results

### Altered cortical functional gradient organization in SCD

To characterize large-scale cortical functional organization, we examined the principal functional gradient derived from resting-state functional connectivity. In both SCD and healthy control (HC) groups, the gradient extended from primary sensory and motor regions to higher-order transmodal association cortices, consistent with a canonical cortical functional hierarchy (Figure 1a).

**Figure 1.**
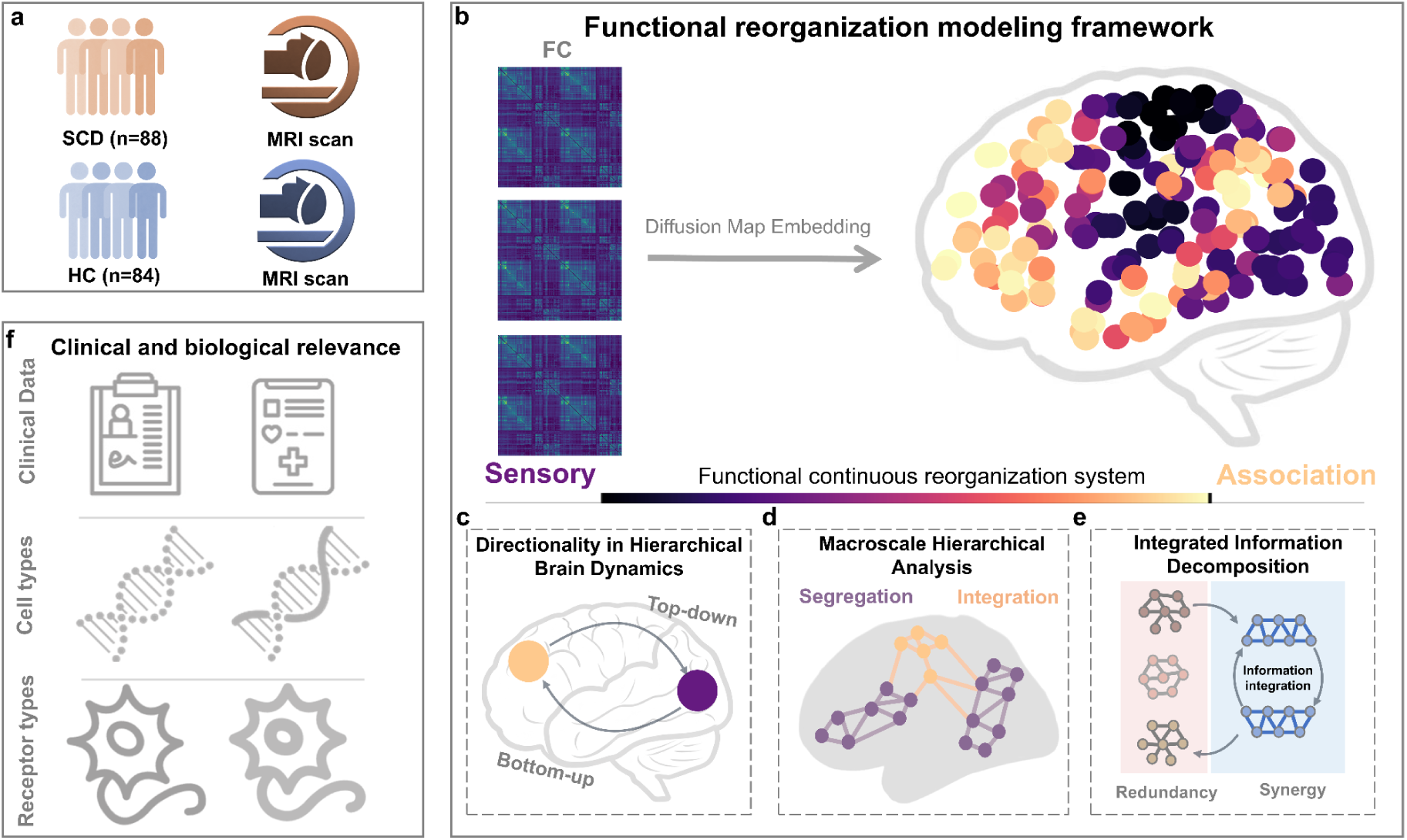
Schematic overview of large-scale reorganization of cortical hierarchy, brain dynamics, and information processing in subjective cognitive decline (SCD). (a) Resting-state fMRI data were obtained from individuals with SCD (n = 88) and healthy controls (HC; n = 84). The right panel illustrates the core analytical pipeline. (b) Whole-brain functional connectivity (FC) matrices were used to construct affinity matrices, and diffusion map embedding was applied to derive the principal functional gradient spanning primary sensory/motor cortices to transmodal association cortices (sensory-to-association axis).The “functional continuous reorganization system” comprises three complementary modules:; (c) directionality in hierarchical brain dynamics, assessing the imbalance between bottom-up (sensory-to-association) and top-down (association-to-sensory) information flow; (d) macroscale hierarchical analysis, quantifying cortex-wide functional integration and segregation using nested spectral partitioning; (e) integrated information decomposition (IID), decomposing distributed information processing into synergistic (synergy) and redundant (redundancy) components and characterizing their network properties (e.g., modularity and efficiency). (f) The imaging-derived measures were linked to clinical characteristics (e.g., symptom duration/transition years, depressive burden) and to molecular–cellular substrates, including neurotransmitter receptor/transporter distributions and cell-type–specific gene-expression profiles, via cross-modal association analyses.

Direct comparison between groups revealed significant regional alterations in functional gradient loadings in SCD relative to HC. These differences were primarily localized to association cortices and survived false discovery rate (FDR) correction *p*_FDR_ < 0.05; Figure 1a). Despite these spatially specific alterations, the overall distribution of functional gradient loadings across the cortex was largely comparable between groups, as indicated by overlapping density distributions (Figure 1b).

In contrast, gradient dispersion—a global measure of variability along the functional hierarchy—was significantly increased in the SCD group compared with HC (*t* = 2.41, *p* < 0.05; Figure 1c). This finding indicates a more heterogeneous and less tightly organized functional hierarchical architecture in SCD.

Importantly, functional gradient alterations were associated with clinical progression within the SCD group. Regions with higher (positive) gradient loadings exhibited a significant positive correlation with transfer years (*r* = 0.38, *p* < 0.05), whereas regions with lower (negative) gradient loadings showed a significant negative correlation (*r* = −0.32, *p* < 0.05; Figure 1d). These results suggest that cortical functional hierarchy undergoes progressive reorganization as subjective cognitive symptoms persist.

### Enhanced bottom-up brain dynamics in SCD and APOE4 carriers

Given the observed alterations in cortical hierarchy, we next examined the directionality of large-scale brain dynamics. Using hierarchical flow analysis, we quantified bottom-up (sensory-to-association) information flow across the cortex (Figure 2a).

**Figure 2.**
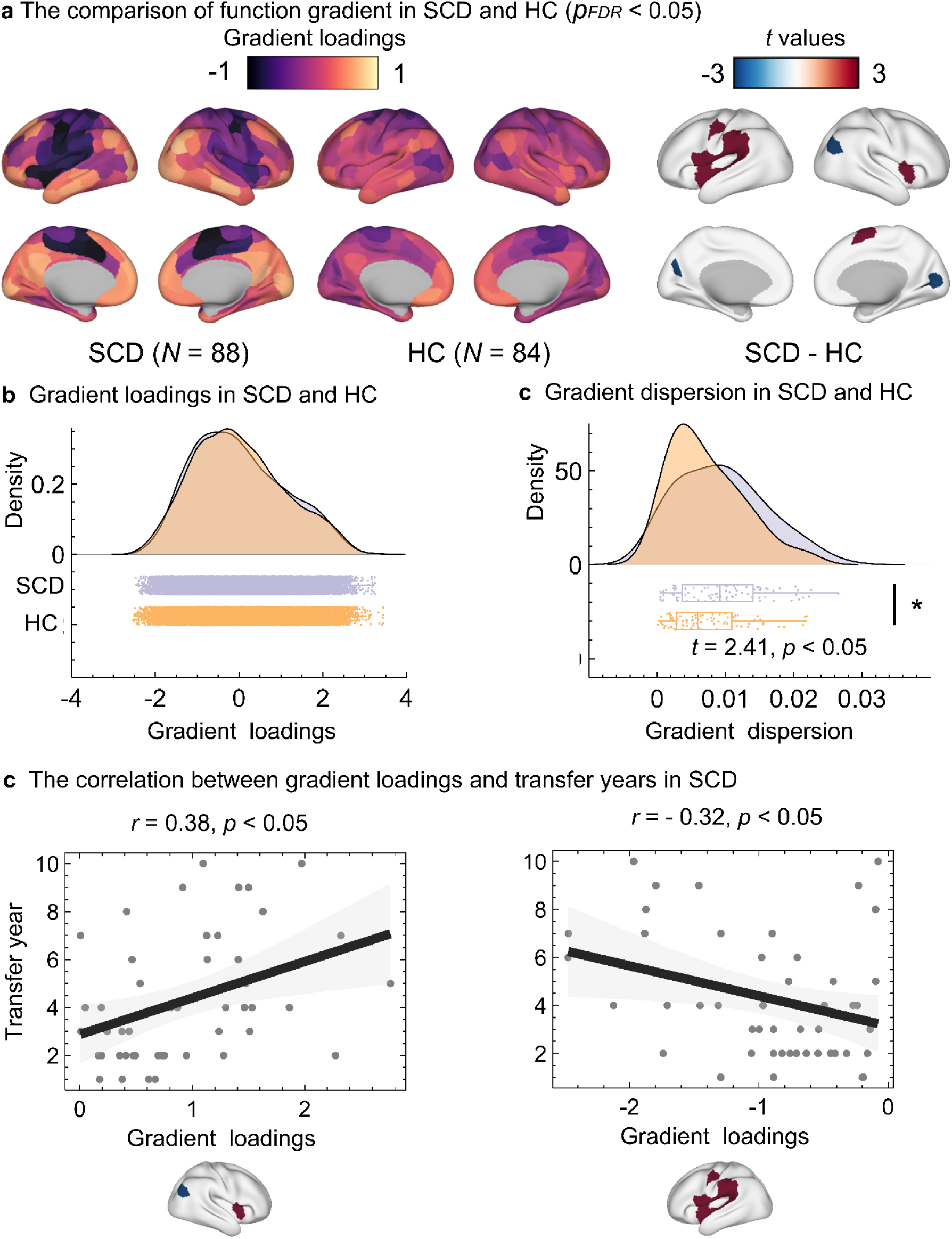
Comparison of the principal functional gradient between SCD and HC. (a) Cortical surface maps illustrating the principal functional gradient in participants with SCD(*N* = 88) and HC(*N* = 84). Gradient loadings are displayed on the cortical surface, spanning from unimodal sensory regions to transmodal association cortices. Between-group differences (SCD − HC) are shown as *t*-value maps and thresholded at *p*_FDR_ < 0.05. (b) Distribution of gradient loadings across cortical regions in SCD and HC. Density plots and scatter distributions indicate largely overlapping global distributions of functional gradient loadings between groups. (c) Gradient dispersion differs significantly between groups, with SCD showing increased dispersion relative to HC (*t* = 2.41, *p* < 0.05), indicating increased heterogeneity of cortical functional hierarchy. (d) Within the SCD group, regional functional gradient loadings are significantly associated with transfer years. Positive gradient loadings show a positive correlation with transfer years (*r* = 0.38, *p* < 0.05), whereas negative gradient loadings show a negative correlation (*r* = −0.32, *p* < 0.05), suggesting progressive reorganization of cortical functional hierarchy with symptom duration.

Participants with SCD exhibited significantly increased bottom-up information flow compared with HC (*t* = 2.18, *p* < 0.05; Figure 2b), indicating a shift toward sensory-driven processing. No corresponding increase in top-down flow was observed, suggesting an imbalance in hierarchical information exchange.

Furthermore, individuals carrying the APOE ε4 allele showed significantly stronger bottom-up dynamics compared with non-carriers (*t* = 2.78, *p* < 0.05; Figure 2c). These findings indicate that enhanced bottom-up information flow is associated both with subjective cognitive symptoms and genetic risk for AD.

### Disrupted functional integration and segregation in SCD

To assess whether altered hierarchical dynamics were accompanied by changes in large-scale network organization, we quantified functional integration and segregation using nested spectral partitioning of whole-brain functional connectomes (Figure 3a).

**Figure 3.**
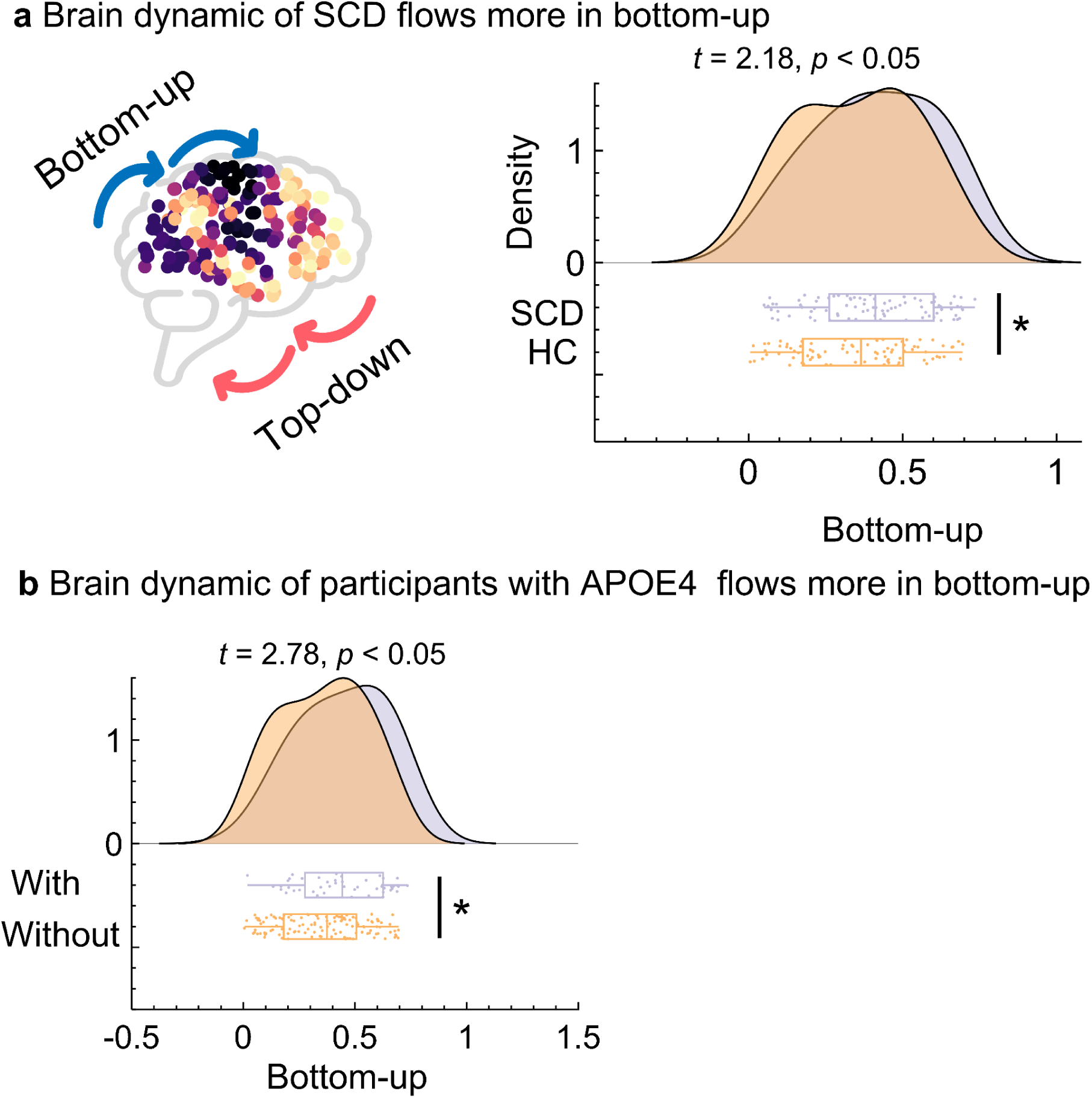
Brain dynamics exhibit enhanced bottom-up information flow in SCD and APOE ε4 carriers. (a) Schematic illustration of bottom-up (sensory-to-association) and top-down (association-to-sensory) information flow across the cortical hierarchy. (b) Quantitative comparison of bottom-up information flow between SCD and HC. Density distributions and boxplots demonstrate significantly increased bottom-up dynamics in SCD relative to HC (*t* = 2.18, *p* < 0.05). (c) Comparison of bottom-up dynamics between participants with and without the APOE ε4 allele. APOE4 carriers exhibit significantly stronger bottom-up information flow compared to non-carriers (*t* = 2.78, *p* < 0.05). Together, these results suggest a shift toward sensory-driven processing and reduced top-down modulation in individuals at risk for cognitive decline.

Compared with HC, the SCD group showed significantly reduced functional integration (*t* = −2.38, *p* < 0.05) alongside significantly increased functional segregation (*t* = −2.27, *p* < 0.05; Figure 3b). This pattern suggests a shift toward more locally segregated but globally less integrated network organization in SCD.

Correlation analyses within the SCD group revealed that functional integration was negatively associated with transfer years (*r* = −0.42, *p* < 0.05; Figure 3c), indicating a progressive decline in global integrative capacity with symptom duration. Functional segregation, however, did not show a meaningful association with transfer years (*r* = −0.012, *p* < 0.05), suggesting relative stability or compensatory maintenance of segregated processing.

### Integrated information decomposition reveals widespread reductions in synergy and redundancy

To further dissect the nature of altered information processing, we quantified synergy and redundancy metrics derived from integrated information decomposition (Figure 4a).

**Figure 4.**
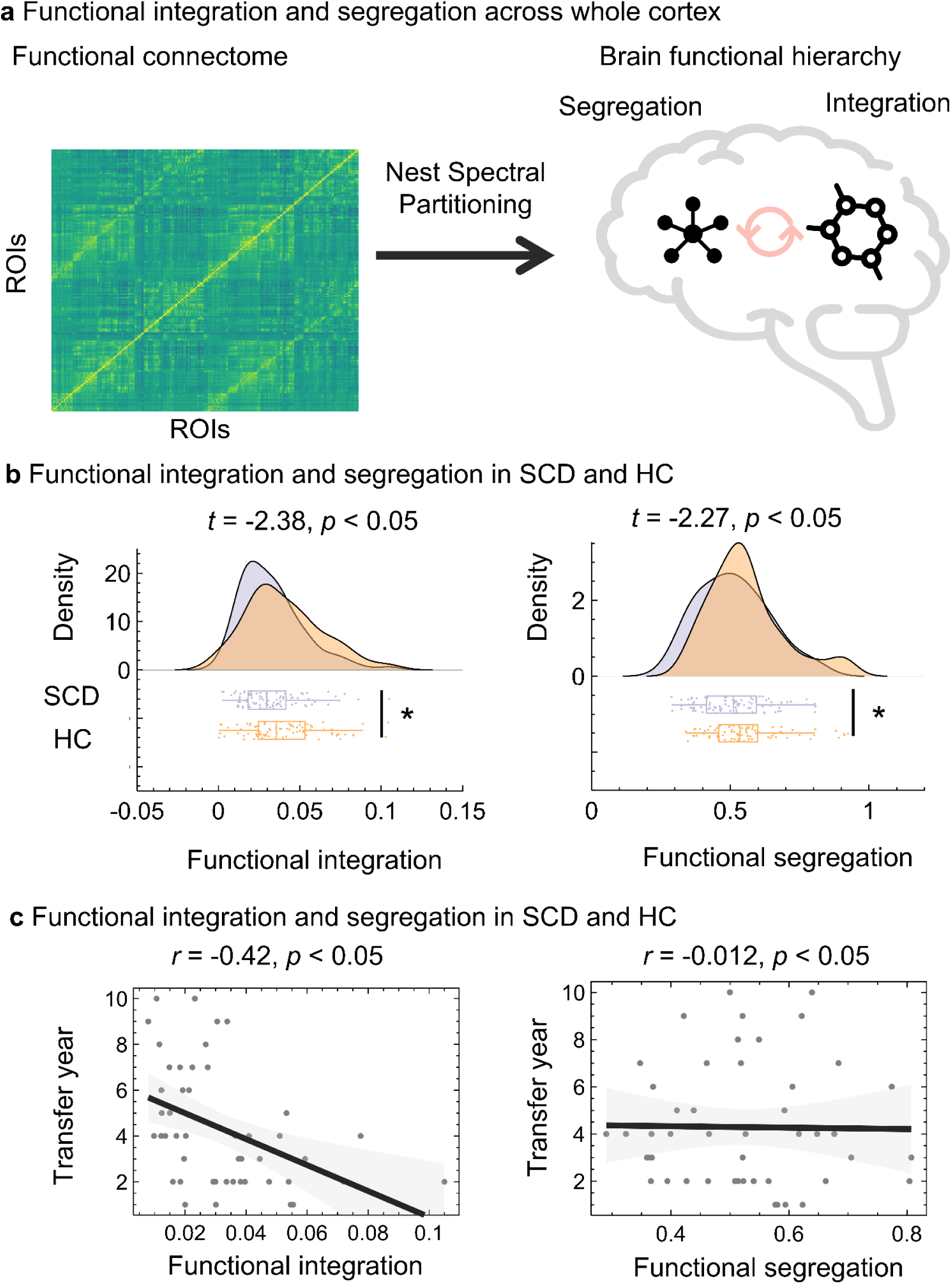
Functional integration and segregation across the whole cortex in SCD. (a) Schematic of functional connectome construction and nested spectral partitioning used to characterize large-scale functional hierarchy, separating integrative and segregative network properties. (b) Group comparisons reveal significantly reduced functional integration (*t* = −2.38, *p* < 0.05) and increased functional segregation (*t* = −2.27, *p* < 0.05) in SCD compared with HC. (c) Within the SCD group, functional integration shows a significant negative correlation with transfer years (*r* = −0.42, *p* < 0.05), whereas functional segregation does not exhibit a significant association (*r* = −0.012, *p* < 0.05). These findings indicate progressive breakdown of large-scale integrative processing with preserved or compensatory segregation in early cognitive decline.

Spatial mapping of the synergy–redundancy gradient revealed marked group differences across association cortices, with significant SCD–HC contrasts surviving FDR correction (*p* FDR < 0.05; Figure 4b). At the global level, participants with SCD showed significantly reduced synergy modularity (*t* = −10.22, *p* < 0.05) and synergy efficiency (*t* = −5.00, *p* < 0.05), indicating impaired integrative information processing (Figure 4c).

In parallel, redundancy-related measures were also significantly reduced in SCD, including redundancy modularity (*t* = −5.41, *p* < 0.05) and redundancy efficiency (*t* = −18.56, *p* < 0.05; Figure 4c). Together, these results suggest a widespread disruption of both synergistic integration and redundant information buffering in early cognitive decline.

Clinical relevance was further supported by significant correlations between information decomposition metrics and SCD symptom severity. Higher Geriatric Depression Scale (GDS) scores were associated with lower synergy modularity (*r* = −0.32, *p* < 0.05), lower synergy efficiency (*r* = −0.23, *p* < 0.05), and lower redundancy efficiency (*r* = −0.22, *p* < 0.05; Figure 4d).

### Molecular and cellular correlates of functional alterations

Finally, we investigated the molecular and cellular substrates underlying the observed functional alterations by correlating cortical functional metrics with neurotransmitter receptor distributions and cell-type–specific gene expression profiles.

Significant spatial correlations (*p*_FDR_ < 0.05) were observed between functional gradient and information decomposition measures and multiple neurotransmitter systems, including serotonergic (5-HT), dopaminergic (D1/D2), GABAergic, glutamatergic (NMDA), and cholinergic (VAChT) markers (Figure 5a). These associations suggest that large-scale functional reorganization in SCD is closely linked to neuromodulatory system architecture.

**Figure 5.**
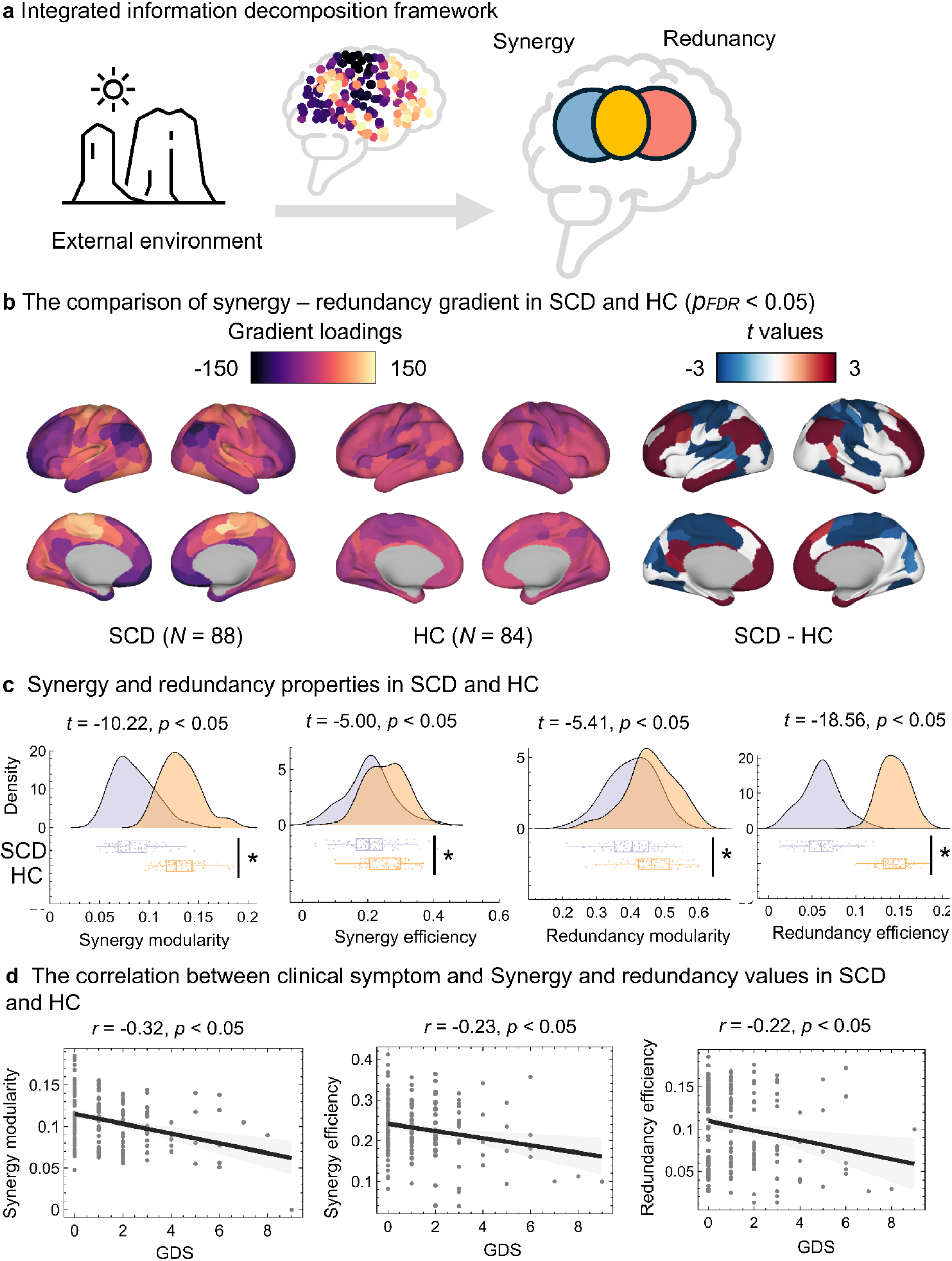
Alterations in synergy and redundancy derived from integrated information decomposition. (a) Conceptual framework illustrating the decomposition of brain information processing into synergistic and redundant components. (b) Cortical maps of the synergy–redundancy gradient in SCD (*N* = 88) and HC (*N* = 84), along with between-group *t*-value maps (SCD − HC), thresholded at *p*_FDR_ < 0.05. (c) Group comparisons of synergy and redundancy metrics. SCD shows significantly reduced synergy modularity (*t* = −10.22, *p* < 0.05) and synergy efficiency (*t* = −5.00, *p* < 0.05), as well as reduced redundancy modularity (*t* = −5.41, *p* < 0.05) and redundancy efficiency (*t* = −18.56, *p* < 0.05), indicating widespread disruption of both integrative and redundant information processing. (d) Correlations between clinical symptoms (Geriatric Depression Scale, GDS) and information decomposition metrics across all participants. Higher GDS scores are associated with lower synergy modularity (*r* = −0.32, *p* < 0.05), synergy efficiency (*r* = −0.23, *p* < 0.05), and redundancy efficiency (*r* = −0.22, *p* < 0.05).

**Figure 6.**
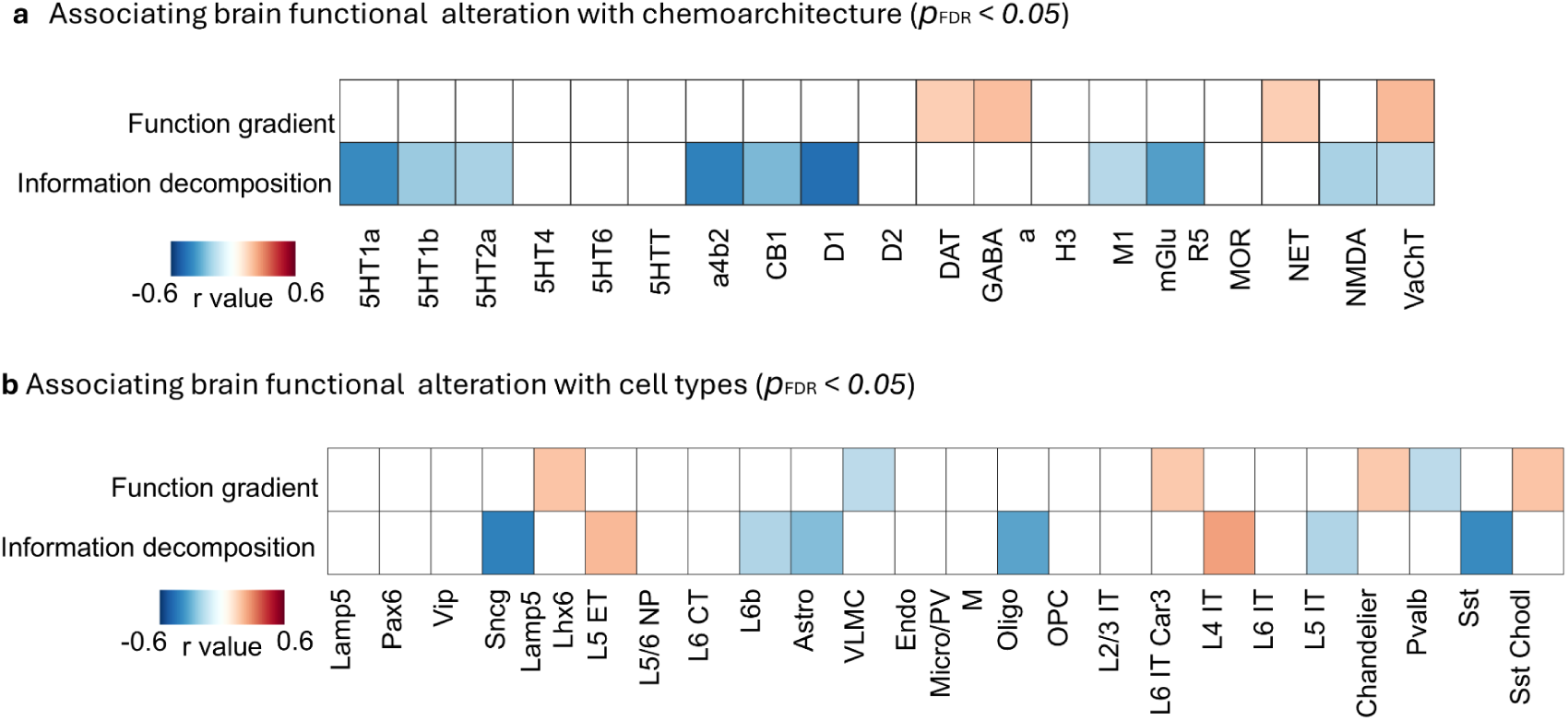
Associations between brain functional alterations, chemoarchitecture, and cell-type expression profiles. (a) Spatial correlations between cortical functional alterations (function gradient and information decomposition metrics) and neurotransmitter receptor/transporter distributions. Significant associations (*p*_FDR_ < 0.05) are observed for serotonergic (5-HT), dopaminergic (D1, D2), GABAergic, glutamatergic (NMDA), and cholinergic (VAChT) systems. (b) Associations between functional alterations and transcriptomic cell-type profiles. Significant correlations (*p*_FDR_ < 0.05) are observed across multiple excitatory, inhibitory, and non-neuronal cell classes, including L2/3 IT, L4 IT, L6 corticothalamic neurons, astrocytes, oligodendrocytes, microglia, and endothelial cells. These results link large-scale functional reorganization in SCD to underlying molecular and cellular substrates.

At the cellular level, functional alterations were significantly associated (*p* FDR < 0.05) with transcriptomic signatures of multiple neuronal and non-neuronal cell types, including excitatory intratelencephalic and corticothalamic neurons, inhibitory interneurons, astrocytes, oligodendrocytes, microglia, and endothelial cells (Figure 5b). This convergence indicates that disrupted cortical information processing in SCD reflects coordinated alterations across molecular, cellular, and systems-level scales.

## Discussion

In this study, we provide a systems-level characterization of early functional brain reorganization in SCD by integrating analyses of cortical hierarchy, hierarchical dynamics, and information-processing architecture. We show that SCD is associated with a loosening of cortical functional hierarchy along the sensory–association axis, a concomitant shift in large-scale brain dynamics toward enhanced bottom-up information flow, and a coordinated reduction in both synergistic and redundant information processing across distributed networks. Together, these findings indicate that subjective cognitive symptoms arise in the context of coherent, multilevel disruptions to how information is organized, propagated, and integrated in the brain, rather than from isolated regional abnormalities. By situating SCD within a unified framework linking spatial hierarchy, temporal dynamics, and computational information processing, our results advance a mechanistic account of SCD as an early functional manifestation of vulnerability along the Alzheimer’s disease continuum.

We first demonstrate that individuals with SCD exhibit significant regional alterations along the principal sensory–association functional gradient, accompanied by increased global gradient dispersion. This finding indicates a more heterogeneous and less stable organization of cortical functional hierarchy. Previous studies have primarily reported functional connectivity or network topology alterations in AD and mild cognitive impairment, whereas investigations at the SCD stage have largely focused on localized connectivity changes or default-mode network abnormalities^28,29^. Recent research reveals significant shifts in default mode network gradient values in MCI and AD patients. This decline, particularly within the posterior default mode network, reflects a disruption or reorganization of the high-level associative cortex’s hierarchical structure during early disease stages^30^. Our results are consistent with emerging evidence from aging and neurodegenerative research suggesting that higher-order association cortices show early vulnerability in hierarchical organization.

Critically, the novel contribution of the present study lies in showing that measurable disorganization of cortical hierarchy is already evident in SCD, even in the absence of objective cognitive impairment. This observation provides a systems-level explanation for the core clinical paradox of SCD—the dissociation between subjective cognitive complaints and preserved neuropsychological performance. Association cortices occupy apex positions within the cortical hierarchy and play a central role in coordinating information flow across sensory and cognitive systems^31^. Increased dispersion of the functional hierarchy likely reflects reduced efficiency and stability of cross-hierarchical coordination, which may degrade perceived cognitive efficiency without necessarily impairing performance on standard cognitive tests. These findings suggest that functional gradient dispersion may serve as a sensitive marker of early functional vulnerability along the AD continuum.

Building upon the observed spatial reorganization of cortical hierarchy, we further show that brain dynamics in SCD are characterized by a systematic shift toward enhanced bottom-up information flow. Efficient cognition depends on a dynamic balance between bottom-up sensory-driven inputs and top-down predictive and regulatory signals. Disruptions of this balance have been reported in aging, depression, and AD^32–34^. Our findings extend this literature by demonstrating that directional alterations in hierarchical dynamics are already detectable at the SCD stage.

The key novelty of this result is that it advances the characterization of SCD-related dysfunction from static spatial organization to dynamic information processing. Enhanced bottom-up flow is biologically plausible in the context of compromised higher-order association cortices: when integrative and predictive functions are weakened, cortical systems may rely disproportionately on incoming sensory signals, reducing the ability to filter noise and uncertainty. This dynamic imbalance offers a mechanistic account of common subjective experiences in SCD, such as heightened sensitivity to environmental stimuli, reduced attentional efficiency, and increased self-monitoring. Importantly, the presence of similar dynamics in individuals carrying genetic risk factors further supports the interpretation that hierarchical dynamic alterations reflect early AD-related vulnerability rather than nonspecific psychological effects.

At the level of information processing, integrated information decomposition revealed widespread reductions in both synergistic and redundant interactions in SCD. Prior work has reported diminished synergistic information in disorders of consciousness, epilepsy, and psychiatric conditions, highlighting impaired integrative computation across distributed brain systems^22,35^. However, evidence at the SCD or preclinical AD stage has been scarce. Our findings not only confirm reduced integrative capacity but also demonstrate a concomitant reduction in redundant information processing.

This dual reduction constitutes a central new insight of the present study. Reduced synergy reflects impaired joint encoding and integration across brain regions, whereas reduced redundancy indicates diminished informational robustness and buffering capacity against noise. Their simultaneous decline suggests that SCD is not merely characterized by reduced processing efficiency, but by a more fundamental reconfiguration of large-scale information architecture. This pattern is theoretically coherent: loosening of hierarchical organization and a shift toward bottom-up dynamics would be expected to culminate in both weakened integrative computation and reduced informational resilience. Thus, information decomposition provides a unifying computational framework that bridges spatial hierarchy and temporal dynamics.

We further show that these functional alterations are significantly associated with symptom duration and depressive burden, indicating close correspondence between large-scale brain reorganization and subjective clinical experience. While previous studies have often attributed SCD symptoms primarily to affective factors, our results suggest that emotional burden may amplify or reveal underlying systems-level functional vulnerability rather than fully account for subjective complaints. Moreover, the spatial correspondence between functional alterations and neurotransmitter receptor distributions as well as cell-type–specific gene expression profiles aligns with growing evidence that neuromodulatory systems and cellular architecture contribute to early functional changes along the AD continuum. Neuromodulatory systems in the early stages of SCD not only regulate local excitation-inhibition balance but also play a central role in shaping the brain’s large-scale functional hierarchical organization. Functional abnormalities in the SCD stage arise not only from neurons but also involve a broad range of non-neuronal cells. Transcriptional perturbations in medial temporal lobe excitatory neurons and corticothalamic neurons may drive declines in memory and integrative functions; dysfunction of inhibitory interneurons disrupts local network synchrony; and altered gene expression in astrocytes, oligodendrocytes, microglia, and endothelial cells suggests that non-cell-autonomous mechanisms—such as neurovascular unit dysregulation, myelin maintenance deficits, and neuroinflammation—also contribute to early functional reorganization.

This multiscale convergence strengthens the interpretation of SCD as a biologically grounded risk state. The observed alterations in cortical hierarchy, dynamics, and information processing are unlikely to represent isolated phenomena; instead, they may reflect the systems-level manifestation of molecular and cellular vulnerability. This perspective provides a principled foundation for integrating functional neuroimaging with molecular imaging and fluid biomarkers in future studies of early AD risk.

## Conclusion

Taken together, our findings delineate a coherent trajectory of early functional reorganization in SCD across spatial, temporal, and informational dimensions. Loosening of cortical functional hierarchy provides a structural substrate for altered dynamics, which in turn give rise to systematic disruptions in information integration and redundancy. This multilevel consistency supports a unified systems-level account of SCD.

Future longitudinal studies will be essential to determine whether these functional signatures predict progression to objective cognitive impairment and to clarify their temporal relationship with molecular pathology. More broadly, our results suggest that integrating cortical hierarchy, brain dynamics, and information-theoretic measures may offer a powerful framework for characterizing early neurodegenerative vulnerability and for developing objective markers of subjective cognitive symptoms.

## Methods

Participants with SCD were identified from the AD Neuroimaging Initiative (ADNI)^36^, the National Alzheimer’s Coordinating Center (NACC)^37^, and the Open Access Series of Imaging Studies–3 (OASIS-3) databases^38^. Use of these datasets was approved by the institutional review boards of the participating research centers, and written informed consent was obtained from all participants. A total of 89 individuals met the inclusion criteria: (1) a baseline diagnosis corresponding to SCD; (2) availability of baseline demographic and clinical information, including age, sex, years of education, body mass index (BMI), hypertension status, smoking and alcohol use, and APOE ε4 genotype; (3) completion of structural MRI (sMRI) and resting-state functional MRI (rs-fMRI) within one year of baseline assessment; and (4) availability of longitudinal clinical follow-up data spanning 10 years from baseline, with outcomes meeting one of the following conditions: (i) progression to mild cognitive impairment (MCI) during the 10-year follow-up period, or (ii) no transition in cognitive status throughout the entire follow-up (i.e., remaining classified as SCD). Detailed diagnostic criteria are provided in the manuals of the respective databases. Briefly, SCD was defined as the presence of self-reported memory decline persisting for at least six months, with normal objective memory performance at baseline. A diagnosis of MCI required objective impairment in at least one cognitive domain, a Mini-Mental State Examination (MMSE) score ≥ 24, or a Montreal Cognitive Assessment (MoCA) score ≤ 26. Exclusion criteria included: (1) neurological disorders such as stroke, cerebral infarction, traumatic brain injury, or brain tumors; (2) psychiatric or psychological conditions including schizophrenia, intellectual disability, or major depressive disorder; and (3) current severe alcohol or substance abuse.

### Participants

Patients with SCD and normal controls were selected from the ADNI, NACC, and OASIS-3 databases. SCD patients (n=88) and healthy controls (n=84) were included based on the following criteria: (1) Baseline status was consistent with the related diagnostic label of either SCD or normal cognition. (2) Availability of baseline demographic and clinical data. (3) Completion of both sMRI and rs-fMRI within one year of baseline. Additionally, SCD patients were required to have at least 10 years of clinical follow-up, with documented progression to mild cognitive impairment within that period or no cognitive conversion throughout the entire follow-up. The detailed diagnostic criteria are described in the manuals of the three databases. Exclusion criteria included: (1) Neurological diseases such as stroke, cerebral infarction, traumatic brain injury, and brain tumors; (2) Psychiatric or psychological disorders such as schizophrenia, intellectual disability, and depression; (3) Current significant alcohol or substance abuse.

### MRI acquisition

Structural MRI data were acquired from three different scanners using a 3D T1-weighted MPRAGE or IR-FSPGR sequence on three different 3T scanners. For the Siemens 3T scanner, acquisition parameters were: repetition time (TR)/echo time (TE) = 2300-2400/2.95-3.16 ms, flip angle = 8-9°, slice thickness = 1.0–1.2 mm. For the GE Medical Systems 3T scanner, parameters were TR/TE = 7.2–7.7/2.9–3.1 ms, flip angle = 11°, slice thickness = 1.0–1.2 mm. For the Philips Medical Systems 3T scanner, parameters were TR/TE = 6.5–6.8/2.9–3.2 ms, flip angle = 11°, slice thickness = 1.0–1.2 mm.

The rs-fMRI data were acquired using standard echo-planar imaging sequences or equivalent protocols from three scanners. For the Siemens 3T scanner, rs-fMRI data were acquired with TR/TE= 2200-3000/27-30 ms, flip angle = 90°, and slice thickness = 3.3–4.4 mm. For the GE Medical Systems 3T scanner, acquisition parameters were TR/TE = 3000/30 ms, flip angle = 90°and slice thickness = 3.4 mm. For the Philips Medical Systems 3T scanner, parameters included TR/TE = 3000/30 ms, flip angle = 80–90°, and slice thickness = 3.3–3.4 mm. Detailed imaging parameters can be obtained from the ADNI (https://adni.loni.usc.edu/), NACC (https://naccdata.org/), and OASIS-3 (https://www.nitrc.org/projects/oasis3/) websites.

### Data preprocessing, quality control, and functional connectome

All raw DICOM images were transformed into the Brain Imaging Data Structure (BIDS) ^39^ format using HeuDiConv (version 0.13.1). Preprocessing of both structural and functional images were carried out with fMRIPrep (version 23.0.2)^40^, which operates on the NumPy^41^ computational framework. Anatomical processing steps encompassed bias field correction and intensity standardization, skull stripping, segmentation of brain tissues, cortical surface reconstruction, and normalization to standard space. Functional image preprocessing included correction for head motion, adjustment for slice acquisition timing, and alignment with the corresponding structural images. The resulting time series were segmented according to the Schaefer 200-region, 7-network atlas. Nuisance signal regression was subsequently applied using Nilearn, following the “simple” denoising approach^42^. This procedure involved temporal high-pass filtering, regression of motion-related parameters and non-neuronal tissue signals, removal of linear trends, and standardization of the time series. The functional magnetic resonance imaging–derived measures were harmonized using ComBat (https://github.com/Jfortin1/ComBatHarmonization), a post-acquisition statistical approach designed to correct site-related batch effects. Finally, individual-level functional connectivity matrices were generated by computing pairwise zero-lag Pearson correlation coefficients between all parcels.

### Functional hierarchy definition

Cortex-wide gradients of the functional connectome were computed using BrainSpace (v0.1.10; https://github.com/MICA-MNI/BrainSpace) with standard parameter settings^43^. In line with previous work, only the strongest 10% of connections for each cortical region were preserved following z-score normalization of the connectivity data. To quantify similarities in regional connectivity profiles, an affinity matrix was generated using a normalized angular kernel. Functional gradients were subsequently identified using diffusion map embedding^44^, a non-linear dimensionality reduction method well suited for high-dimensional connectomic data. This procedure was applied at the individual level to obtain participant-specific gradient representations. Finally, individual gradients were aligned to reference gradients estimated from the group-averaged functional connectome to ensure cross-subject correspondence.

### Dispersion of Functional reorganization in SCD and HC

We defined global dispersion as the Euclidean distance between each individual brain region and the centroid of the manifold. This metric, equivalent to eccentricity, provides a quantitative index of whole-brain functional integration and segregation. Brain regions with higher global dispersion values exhibit greater functional segregation, whereas regions with lower values demonstrate stronger functional integration with nearby areas. Differences in gradient dispersion between individuals with SCD and HC were assessed using two-sample t tests.

### Cognition decoding for functional reorganization

We then further estimated the cognition relevance of functional reorganization patterns among various adolescent subtypes using Neurosynth^45^. We used twenty cognition maps from previous studies^46^, which represented sensory functions and high-order cognition. Then we correlated them with functional gradient difference maps between SCD and HC via Pearson correlation.

### Top-down and bottom-up information flow in SCD and HC

We additionally examined the directionality of information transfer along the sensory–association continuum. Information transmission from association cortices to sensory areas was operationalized as top-down signaling, whereas the reverse direction, from sensory to association regions, was defined as bottom-up signaling^18^. Directed information flow was quantified using regression dynamic causal modeling (rDCM)^47,48^ in combination with graph-theoretical metrics. Subsequently, we computed the ratio of mean outward connectivity strength in sensory regions relative to that in association regions. Values of this ratio below unity were interpreted as reflecting predominant top-down information flow, whereas values exceeding one indicated dominance of bottom-up signaling. Comparisons of flow ratios across SCD and healthy control participants were conducted using independent two sample ttest, while controlling for head motion, age, handedness, and sex as covariates.

### Functional integration and separation of cortex-wide connectome in SCD and HC

To characterize cortex-wide functional integration and segregation, we employed the Nested Spectral Partitioning (NSP) approach ^49,50^, which leverages an eigenmode-based framework to identify hierarchical modular organization and to quantify the brain’s capacity for functional segregation and integration within functional connectivity (FC) networks. Unlike conventional clustering or modularity-optimization techniques, the NSP method is grounded in a physical principle whereby brain regions sharing the same eigenvector sign are interpreted as being co-activated, whereas regions with opposite signs are considered to exhibit antagonistic activation patterns ^49^.

Specifically, spectral graph theory was applied to compute the Laplacian matrix derived from the whole-brain FC matrix. The resulting eigenvalues and eigenvectors were then used to generate an initial partition of the functional network. This spectral decomposition yields a division that minimizes the number of cut edges between regions, thereby defining the primary functional modules. Following this initial segmentation, each module undergoes further subdivision in a recursive and hierarchical manner. This nested partitioning continues until a predefined spatial scale is reached or alternative stopping criteria are satisfied.

At each hierarchical level, intra-subnetwork connectivity strength is calculated to index functional segregation, whereas inter-subnetwork connectivity strength is computed to represent functional integration^50^. Together, these measures provide a multiscale quantification of segregation and integration properties across the brain’s functional architecture. Although NSP-derived metrics are known to be sensitive to the temporal length of the time series used for FC estimation, this potential confound was controlled in the present study, as all participants underwent resting-state data acquisition with identical scan durations. Finally, segregation and integration coefficients of whole-brain functional connectivity were calculated for each participant, and group differences between the disease group and healthy controls were assessed using two-sample *t*-tests, with age and sex included as covariates in the statistical model.

### Synergy and redundancy: integrated information decomposition

The Integrated Information Decomposition (IID)^23^ framework integrates core principles from Integrated Information Theory (IIT) and Partial Information Decomposition (PID) to characterize patterns of information transfer within neural systems^51^. Extending Shannon’s mutual information, IID quantifies statistical dependence between variables and further decomposes the information contributed by two source processes about a target into unique, redundant, and synergistic components^25^. Synergistic information captures contributions that emerge only through the joint interaction of both sources, whereas redundant information reflects shared contributions that each source can independently provide. By enabling this decomposition, the IID framework offers a principled means to examine how multiple brain regions jointly shape future neural states, thereby clarifying the complementary roles of synergy and redundancy in large-scale neural coordination. To overcome limitations of the original IIT formulation of integrated information (Φ), which may produce negative estimates in systems dominated by redundancy, Mediano and colleagues proposed a revised metric (Φ_R) that explicitly accounts for redundant information^52^. This reformulation guarantees non-negative values and yields a more stable and interpretable estimate of integrated information by emphasizing synergistic interactions^53^. Within this framework, brain regions can be conceptualized as “gateways,” which promote integration through synergistic interactions, or as “broadcasters,” which facilitate the dissemination of redundant information across the network.

Applying the IID framework to time series data parcellated using the Schaefer 200 × 7 atlas, we computed, for each participant, a 200 × 200 synergy matrix and a corresponding 200 × 200 redundancy matrix, representing pairwise synergistic and redundant relationships among cortical regions.

### Gradient of redundancy-to-synergy relative importance

We next assessed the relative involvement of each region of interest (ROI) in synergistic versus redundant interactions. For both synergy and redundancy networks, nodal strength was calculated as the sum of all connections for each region based on the group-averaged matrices. All 200 regions were subsequently ranked according to nodal strength, with higher ranks assigned to regions exhibiting stronger connectivity within each network^25^. By subtracting the redundancy rank from the synergy rank for each region, we derived a continuous gradient score spanning negative to positive values. Negative scores denote a predominance of redundancy-related connectivity, whereas positive scores indicate greater engagement in synergistic interactions. This gradient captures regional differences in the balance between redundancy and synergy, thereby illuminating how individual brain areas preferentially support either distributed information sharing or integrative processing within the broader network architecture.

### Graph-theoretic profiles for synergy and redundancy properties

To further delineate the organizational properties of synergy and redundancy networks, we quantified modularity and efficiency, two fundamental graph-theoretical metrics. Modularity reflects the extent to which a network segregates into densely interconnected modules, providing insight into its functional organization. Within synergy networks, elevated modularity suggests efficient large-scale integration supporting global information processing, whereas in redundancy networks, higher modularity emphasizes localized, parallel information transmission that may enhance functional robustness. Conversely, reduced modularity may indicate disrupted network segregation, potentially contributing to impairments in cognitive or affective regulation.

Network efficiency was assessed at both local and global scales. Local efficiency indexes short-range information transfer and supports specialized regional processing, while global efficiency reflects the capacity for long-distance communication and integrative processing across the brain. In the context of SCD, reductions in global efficiency may signify compromised network integration, whereas increases in local efficiency could reflect compensatory reorganization^25^.

Accordingly, modularity and efficiency were computed for both synergy and redundancy matrices at the individual level. Group differences among SCD and healthy controls were evaluated using two sample ttest, controlling for head motion, age, handedness, and sex.

### Molecular mechanism of functional reorganization of SCD

In the present work, cortical magnetic gradient patterns were spatially associated with 24 previously published cell-type–specific maps. Molecular expression signatures for each cellular class were derived from single-nucleus Drop-seq (snDrop-seq) datasets reported in earlier work. Cell-type abundance across the cortex was subsequently inferred by deconvolving microarray expression data from the Allen Human Brain Atlas. The resulting set comprised 24 distinct cell populations, including excitatory and inhibitory neuronal subtypes as well as non-neuronal cell classes: Lamp5, Pax6, Vip, Sncg, Lamp5 Lhx6, L5 ET, L5/6 NP, L6 CT, L6b, astrocytes, VLMC, endothelial cells, microglia/perivascular macrophages, oligodendrocytes, oligodendrocyte precursor cells, L2/3 IT, L6 IT Car3, L4 IT, L6 IT, L5 IT, chandelier cells, Pvalb, Sst, and Sst Chodl.

Neurotransmitter receptor and transporter distributions were quantified using positron emission tomography (PET)–derived maps encompassing 19 targets across nine major neurotransmitter systems. These datasets, recently compiled and released by Hansen and colleagues (https://github.com/netneurolab/hansen_receptors)^54^. include markers for dopamine (D1, D2, DAT), norepinephrine (NET), serotonin (5-HT1a, 5-HT1b, 5-HT2a, 5-HT4, 5-HT6, 5-HTT), acetylcholine (α4β2, M1, VAChT), glutamate (mGluR5, NMDA), γ-aminobutyric acid (GABAa/bz), histamine (H3), cannabinoid (CB1), and opioid (MOR) systems^55^. All volumetric PET images were spatially normalized to the MNI 152 nonlinear 2009 asymmetric template (version c) and averaged across participants within each study. The resulting images were then parcellated according to the Schaefer 400-region atlas. For receptors or transporters represented by multiple PET tracers (specifically 5-HT1b, D2, and VAChT), weighted averaging was applied to generate a single representative map for each target.

### Null model

To characterize spatial correspondences between common- and SCD network configurations and other neurobiological features, we employed a spatially informed null modeling framework. This approach was designed to disrupt the correspondence between two cortical maps while preserving their intrinsic spatial autocorrelation structure^56^. Initially, receptor distribution maps were randomly permuted and their associations with network patterns were recalculated. These spatial coordinates were then used to generate null distributions through repeated random rotations, followed by reassignment of node values to the nearest cortical parcel. This process was iterated 1,000 times to establish stable null distributions. Importantly, rotations were first applied to one cerebral hemisphere and subsequently mirrored to the opposite hemisphere to maintain interhemispheric symmetry. Statistical significance was defined using the *95th* percentile of the resulting null distributions derived from both spatial and temporal models. This stringent procedure enabled the identification of meaningful topographic relationships while effectively accounting for spatial dependencies inherent in cortical data.

## Data Availability

The clinical data used in this study involves patient privacy concerns. Therefore, access to the data can be granted upon reasonable request to the corresponding authors. Allen Human Brain Atlas (AHBA; http://human.brain-map.org/), Neuromap (https://netneurolab.github.io/neuromaps/usage.html) could be downloaded online.

## Code Availability

All codes used in this study were available (https://github.com/Laoma29/adolescence-MDD-subtyping-projects). Brainspace (https://brainspace.readthedocs.io/en/latest/) and ENIGMA toolbox (https://enigma-toolbox.readthedocs.io/en/latest/pages.html) could be downloaded online. Data for this study were obtained from the ADNI (https://adni.loni.usc.edu/), NACC (https://naccdata.org/), and OASIS-3 (https://www.nitrc.org/projects/oasis3/) websites. All data were publicly available from these databases.

## Data Availability

All data produced in the present study are available upon reasonable request to the authors

## Acknowledgments

Xiaobo Liu is supported by the China Scholarship Council.

## Author Contributions Statement

X.B.L., J.B.F., and C.M.L. designed research; X.B.L. and X.Y.L. analyzed the data; X.B.L. and X.Y.L. draw the figures; X.B.L., J.B.F., and C.M.L. wrote the first draft of the manuscript; X.B.L., J.B.F., X.Y.L., Z.Y.H., Q.B.Z. and C.M.L. edited the manuscript.

## Supplementary Information

**Supplementary Figure 1.**
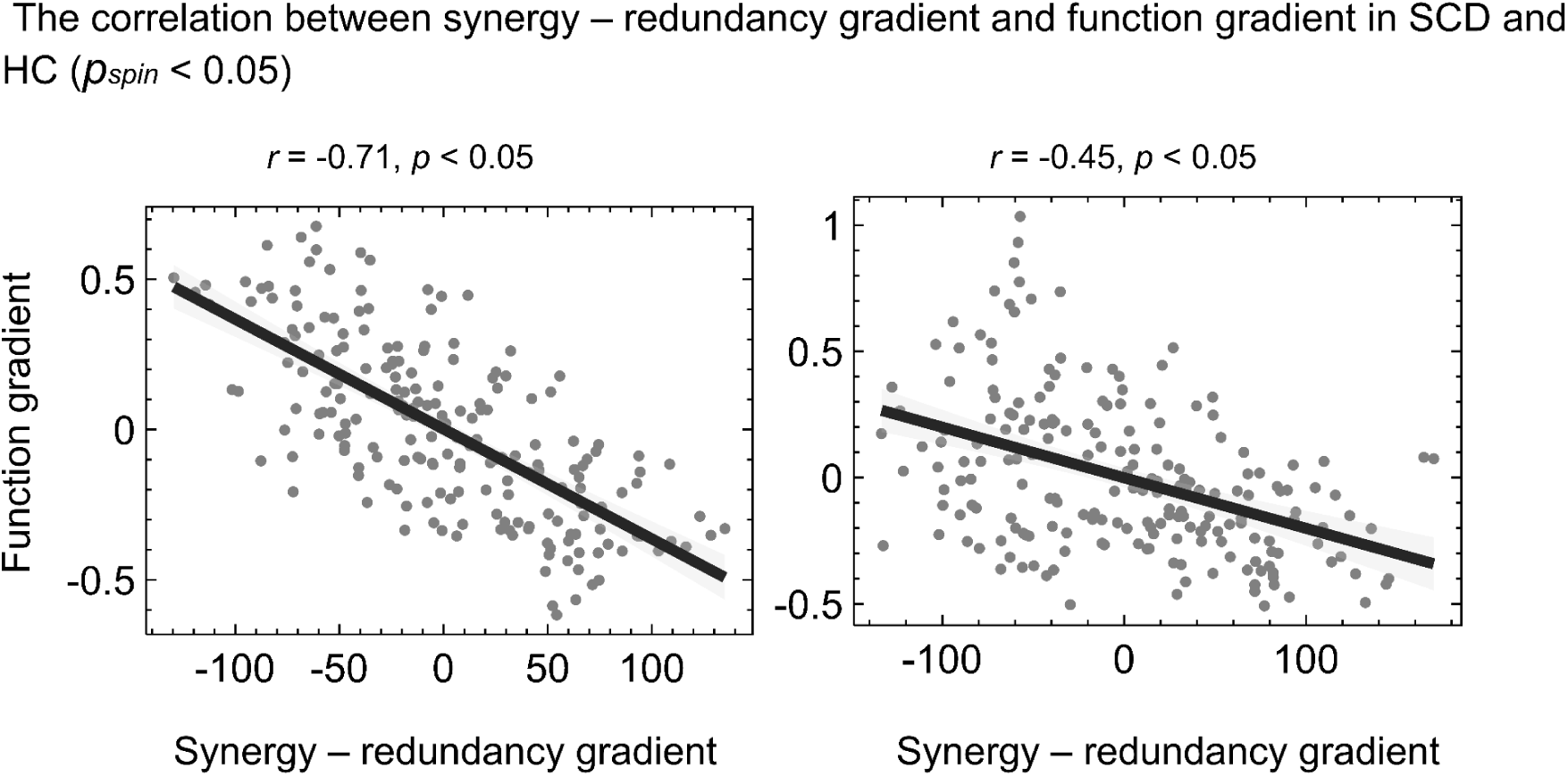
Correlation between the synergy–redundancy gradient and the functional gradient in SCD and healthy controls. Scatter plots depict the spatial association between the synergy–redundancy gradient and the principal functional gradient across cortical regions in individuals with subjective cognitive decline (SCD; left) and healthy controls (HC; right). Each dot represents a cortical parcel. Solid black lines indicate linear regression fits, and shaded regions denote 95% confidence intervals. Significant negative correlations were observed in both groups (spin-test–corrected *p* < 0.05), with a stronger correlation in the SCD group (*r* = −0.71) compared with HC (*r* = −0.45).

**Supplementary Figure 2.**
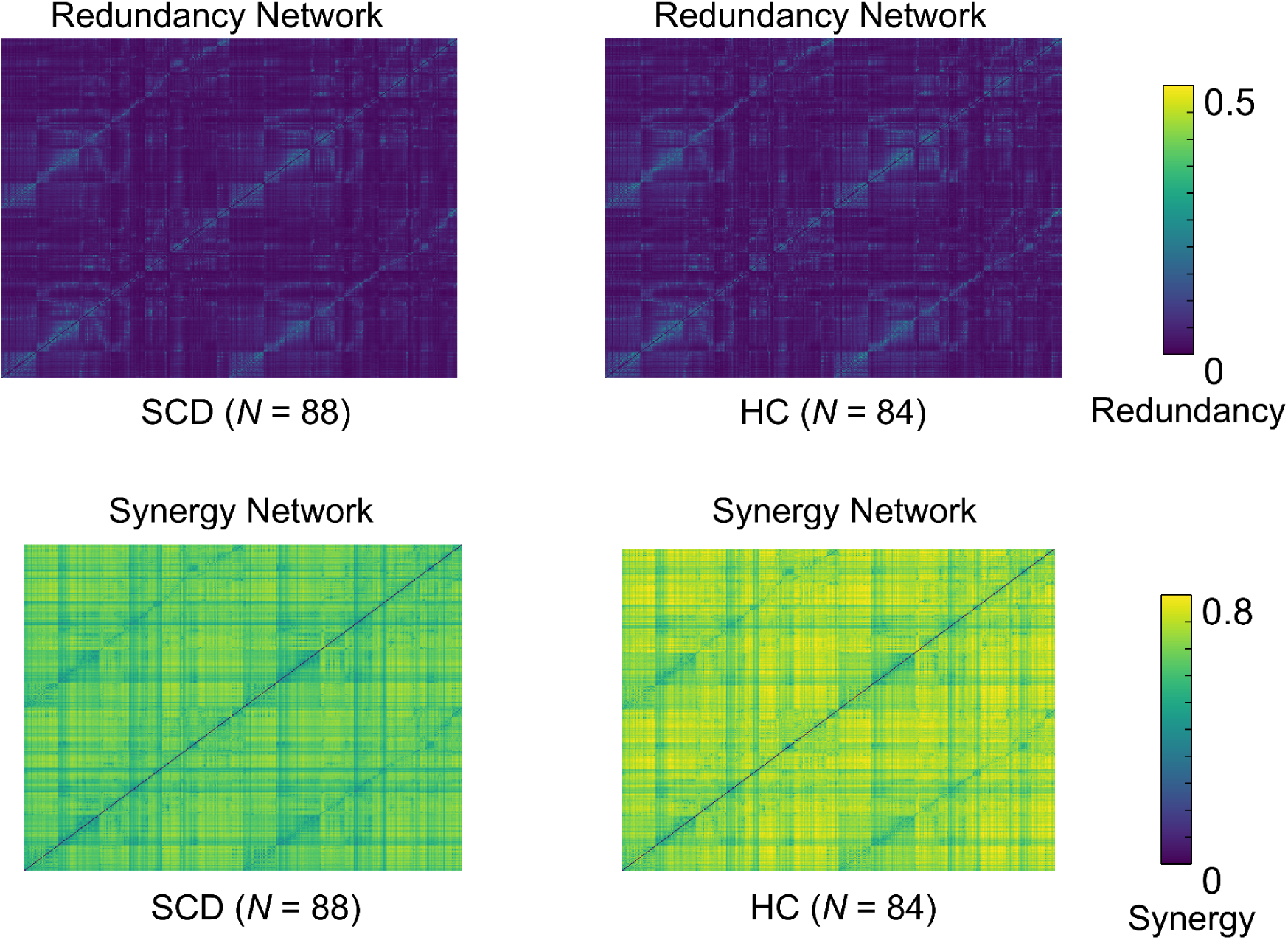
Group-averaged redundancy networks in SCD and healthy controls. Group-level redundancy networks for the SCD group (left; *N* = 88) and healthy controls (HC; right; *N* = 84) are shown as region-by-region matrices. Each matrix element represents the magnitude of redundant information shared between pairs of cortical regions. Warmer colors indicate higher redundancy values, reflecting greater overlap in information contribution across regions.

**Supplementary Table 1.** Demographic information for datasets used in this study. (HAMD, Hamilton Depression Rating Scale; HAMA, Hamilton Anxiety Rating Scale)

|  | Group | Number | Age | Sex (F/M) | Education, year | APOE4 (with/without) | GDS | Mean FD |
| --- | --- | --- | --- | --- | --- | --- | --- | --- |
| **ADNI Datasets.** | SCD | 16 | 75.93<br>±6.46 | 8/8 | 16.75<br>±2.62 | 4/12 | 1.31 ±1.40 | 0.12±0.021 |
|  | HC | 29 | 73.46<br>±5.69 | 12/17 | 16.35<br>±2.35 | 10/19 | 1.35 ±1.80 | 0.11±0.021 |
|  | P values |  | 0.191 | 0.578 | 0.598 | 0.748 | 0.951 | 0.21 |
| **NACC Datasets** | SCD | 3 | 71.33<br>±3.51 | 2/1 | 17.00<br>±2.65 | 2/1 | 1.33 ±0.58 | 0.13±0.041 |
|  | HC | 12 | 76.75<br>±5.51 | 10/2 | 15.83<br>±1.80 | 0/12 | 0.83 ±1.19 | 0.12±0.020 |
|  | P values |  | 0.134 | 0.517 | 0.372 | 0.027 | 0.502 | 0.80 |
| **OASIS-3 Datasets.** | SCD | 69 | 72.25<br>±6.36 | 40/29 | 15.51<br>±2.76 | 18/51 | 1.93 ±1.99 | 0.12±0.025 |
|  | HC | 43 | 69.94<br>±4.90 | 26/17 | 15.91<br>±2.45 | 12/31 | 0.81 ±1.03 | 0.11±0.024 |
|  | P values (two sample *t*-test, two side) |  | 0.038 | 0.794 | 0.438 | 0.833 | 0.002 | 0.31 |

